# Cost of tuberculosis-related aeromedical retrievals in the Torres Strait, Australia

**DOI:** 10.1101/2022.01.13.22269264

**Authors:** J’Belle Foster, Daniel Judge, Diana Mendez, Ben J. Marais, Dunstan Peniyamina, Emma S. McBryde

**Affiliations:** Australian Institute of Tropical Health and Medicine, Townsville, Queensland, Australia; James Cook University College of Medicine and Dentistry, Townsville, Queensland, Australia; Torres and Cape Tuberculosis Control Unit, Thursday Island, Queensland, Australia; James Cook University Division of Tropical Health and Medicine, Townsville, Queensland Australia; Sydney Institute for Infectious Diseases and Biosecurity, Westmead, New South Wales, Australia; Cairns Tropical Public Health Service, Cairns, Queensland, Australia

## Abstract

Tuberculosis (TB) remains a disease of public health significance at the Australia / Papua New Guinea (PNG) international border. In the remote Torres Strait Islands, aeromedical evacuation is a necessary but costly component of TB management and patients with critical care needs require support to prevent onward TB transmission. A detailed costing of an exemplar TB patient from PNG who presented to a Queensland Health facility in the Torres Strait and required urgent aeromedical evacuation was performed. Data were drawn from patient charts, financial and clinical information systems used within Queensland Health and the Torres and Cape Hospital and Health Service. The total cost of aeromedical evacuation was AUD 124,280; 54% of the cost was attributed to travel. Between 2016 and 2019, 19 patients diagnosed with TB were medically evacuated from an outer Torres Strait Island with a median length of hospital stay of 57 days. Aeromedical evacuation and medical management costs require adequate budget allocation.

## Introduction

The Papua New Guinea (PNG) / Torres Strait border area is potentially a major source of drug susceptible, mono-resistant, drug-resistant and multi-drug resistant TB (MDR-TB) into Australia [1]. At the closest point, the Torres Strait Islands in Queensland are less than 5kms from PNG’s Western Province [2]. Cross border dynamics and current TB prevention and control efforts are largely attributable to geographical, regional, political and administrative complexities and structures [3]. Prior to international border closures in March 2020 due to the COVID-19 pandemic [4], the Torres Strait Treaty allowed residents of 13 Treaty villages in the Western Province of PNG and 14 communities of the Torres Strait Islands in Australia to cross the international border for traditional purposes without health checks, visas or passports [5] (Figure 1). This freedom of movement coupled with the conservative TB incidence rate of 674 cases per 100,000 population in the Western Province of PNG [6], places the Indigenous population of the Torres Strait Islands of Queensland, Australia particularly vulnerable to TB transmission [7].

**Figure 1.**
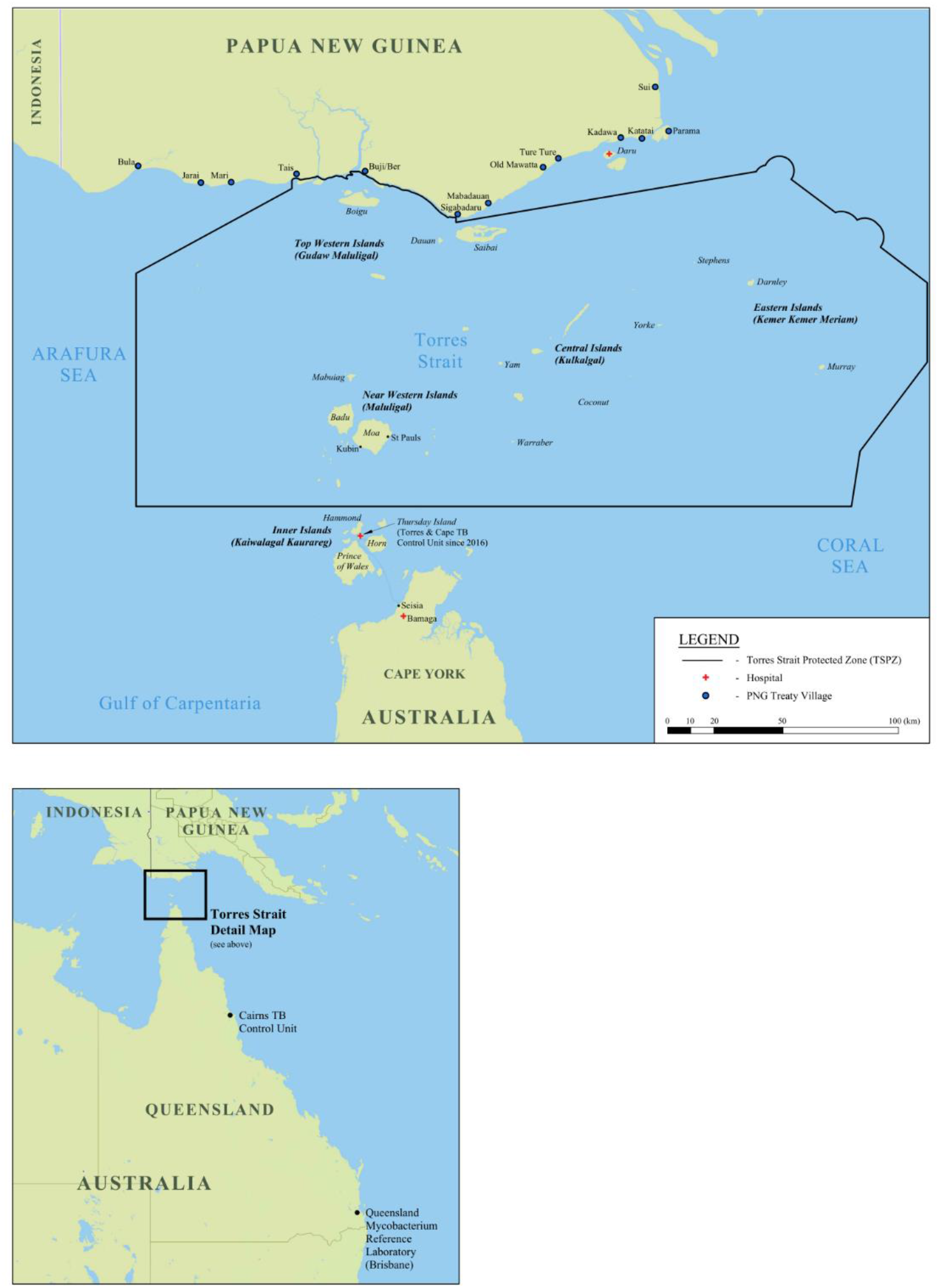
Map of A) the Torres Strait / Papua New Guinea cross-border region and B) relevant tuberculosis referral centres in Queensland, Australia. CC BY 4.01

PNG Treaty villages are among the poorest and most remote in PNG and lack access to clean running water and electricity [2]. Health service provision in the PNG Treaty villages is poor and in 2018, seven basic medical units and one health centre were frequently unstaffed or unstocked with basic medical supplies. The closest major health facility from the Treaty villages in the Western Province of PNG, is Daru General Hospital, located at least two hours away by personal boats powered by small outboard motors [2]. Hence, PNG residents frequently cross the narrow stretch of water separating the two countries to attend well-resourced Queensland Health Primary Health facilities located in the Torres Strait Islands in Australia [3].

Aeromedical evacuation is a necessary component of Queensland Health’s obligation to provide critical health care services to anyone presenting to Torres and Cape Hospital and Health Service (TCHHS) primary health centres (PHCs) in the Torres Strait Protected Zone (TSPZ) including residents of PNG that live adjacent to the Torres Strait Islands (Figure 1). In recognition of health services provided to PNG nationals within the Queensland Health system, funding provided through the Commonwealth’s ‘Managing Torres Strait / Papua New Guinea Cross Border Health Issues (Schedule B) National Partnership Agreement’, was AUD 18.9 million, between 1 July 2016 to 30 June 2020, averaging AUD 4.7 million per year [8]. This study describes detailed costing of the medical evacuation and medical management of one PNG national patient diagnosed with pulmonary TB who presented to Boigu Island in 2019 and who was subsequently admitted to both Thursday Island and Cairns Hospitals (Figure 1).

Medicare is Australia’s health insurance scheme which provides free or low-cost access for all Australians to public hospitals, other medical services and medication [9]. Typically, overseas visitors whose countries do not have a Reciprocal Health Care Agreement with Australia are considered to be Medicare ineligible [10]. Medicare ineligible patients are responsible for fees and charges associated with both outpatient and inpatient care in Queensland [10]. As PNG does not have a Reciprocal Health

Care Agreement with Australia [11], PNG nationals without a Medicare card who enter Australia via a designated port, must pay for health services received. The exception to this rule is for PNG nationals who enter the Australian health care system via the TSPZ [12]. While these patients are still Medicare ineligible, costs incurred for managing and treating them are absorbed by Queensland Health Hospital and Health Services (HHSs), with partial funding to offset costs provided by the Australian Government Department of Health. This, in addition to remoteness and travel, are some of the reasons that the cost of health care per capita / per service rendered, is higher for HHSs in Far North Queensland than is typical across funding structures in other HHSs in Queensland [12].

The Australian Government provides important and strategic funding to manage the delivery of health services to PNG nationals which specifically includes but is not limited to the management of TB patients [8] to address transmission risk from PNG to Australia [13]. This funding is not intended to be used for PNG patients who have arrived in Australia with a passport at an authorised port, rather, it is designated for HHSs to provide healthcare to PNG nationals entering via the TSPZ. At no ‘out of pocket’ expense for PNG patients, funding includes but is not limited to: hospital admissions, outpatient services, management of TB patients, pathology, pharmaceuticals and patient transport [8]. Although Schedule B of the National Partnership Agreement states that funding for health services rendered includes ‘*patient transport (such as medical evacuations)*’ [8], the Commonwealth funding does not cover the majority of expenses incurred by the services undertaking or supporting aeromedical evacuations such as Aeromedical Retrieval and Disaster Management Branch (ARDMB)-supported aeromedical evacuation, and these additional costs are borne by Queensland Health. This Federal funding allocated to managing PNG nationals seeking health care via the TSPZ is absorbed into the total funding pool for all health services rendered in HHSs.

## Materials and Methods

The journey of a PNG national patient who presented to Boigu Island PHC in a critical condition and diagnosed with fully-susceptible pulmonary TB, was selected to cost a typical aeromedical retrieval case of TB. The case was considered typical (without excessive additional costs) as the patient was not accompanied by an escort and did not have drug-resistant TB or comorbidities which may have resulted in an extended hospital admission.

Details of the clinical presentation and pharmaceuticals, pathology, imaging and procedures ordered were reviewed in the TCHHS electronic medical record system, Best Practice, and in the TB patient information database used by the Torres and Cape TB Control Unit. Pathology and imaging services provided were cross checked against Queensland’s laboratory results database AUSLAB, imaging databases Enterprise PACS and Merlin Web, and state-wide health service database The Viewer [14] (Appendix A). The Viewer and Best Practice were also accessed to ascertain the length of hospital stay for patients who were medically evacuated.

Aeromedical evacuation (Boigu Island to Thursday Island; Thursday Island to Horn Island and Horn Island to Cairns) costs to support this patient were sourced from the ARDMB. The ARDMB centrally manage funding and contracts with the Queensland Department of Health and Queensland Ambulance Service (QAS). QAS fund associated paramedic support when required during aeromedical evacuations in TCHHS. Both ARDMB and QAS sit within Queensland Health.

Clinicians within TCHHS will contact Retrieval Services Queensland (RSQ) with requests for aeromedical retrieval. RSQ are part of the ARDMB and centrally coordinate triaging, and the dispatch of paramedic support and aircraft including the rescue helicopter to respond to medical events requiring aeromedical evacuation [15,16]. The Royal Flying Doctors Service (RFDS) provide additional support for inter-facility transfers between Horn Island and Cairns on fixed-wing aircraft. Costs of RFDS interfacility transfer flights are funded by ARDMB through the Queensland Department of Health as part of a state-wide contract to provide aeromedical support to Queensland communities (Appendix A). ARDMB will then recoup a proportion of costs from TCHHS. Costs were expressed in Australian dollars. See Appendix A for extended methodology.

Written consent from the patient was obtained for this study. Ethical approval was granted by the Far North Queensland Human Research Ethics Committee (HREC) (HREC/17/QCH/74-1157), and the Chair of James Cook University HREC, (H7380). Public Health Act authorisation (QCH/36155 – 1157) was also obtained to access patient data. Written approval was separately obtained from the Queensland ARDMB Research Committee, and Queensland Ambulance Service Torres and Cape York Local Ambulance Service Network.

## Results

Table 1 shows that the cost to medically evacuate and medically manage one PNG national with uncomplicated fully-susceptible pulmonary TB from Boigu Island in 2019 was AUD 124,280. The total cost to medically evacuate the patient was AUD 58,029, with the remainder costs attributed to pathology, outpatient assessment, inpatient stay, pharmaceuticals and other travel. The Rescue 700 flight from Boigu Island to Thursday Island and then from Thursday Island to Horn Island Airport comprised the single biggest expense (AUD 48,900), followed by in-patient care in Cairns Hospital (AUD 45,760).

**Table 1.** Cost of managing a Papua New Guinea patient with drug-susceptible tuberculosis (TB) requiring aeromedical retrieval from Boigu Island to the Australian health system in 2019^2^.

| Activity | Funded via Hospital and Health Services (HHSs) |  |  |  | Funded via Queensland Health but external to HHSs |  | Total |
| --- | --- | --- | --- | --- | --- | --- | --- |
|  | Boigu Island Primary Health Centre (TCHHS) | Thursday Island Hospital (TCHHS) | Cairns Hospital (CHHS) | Torres and Cape TB Control Unit (TCHHS) | Queensland Aeromedical Retrieval and Disaster Management Branch (ARDMB) | Queensland Ambulance Service (QAS) |  |
| Single outpatient clinic visit<sup>†</sup> | 1,141 |  |  | 282 |  |  | 1,423 |
| Outpatient pathology | 1,095 |  |  |  |  |  | 1,095 |
| Aeromedical retrieval |  | 6,273<sup>§</sup> |  |  | 2,457<sup>§</sup><br>48,900<sup>‡</sup> | 399<sup>††</sup> | 58,029 |
| Inpatient hospital stay<sup>¶</sup> |  | 8,005 | 45,760 |  |  |  | 53,765 |
| Other travel<sup>¶¶</sup> |  | 8996 |  |  |  |  | 8996 |
| Pharmaceuticals | 35 | 202 | 735 |  |  |  | 972 |
| **TOTAL** | **2,271** | **23,476** | **46,495** | **282** | **51,357** | **399** | **124,280** |

<sup>2</sup> † Category 2 Emergency
¶ Includes costs associated with meals, wages, cleaning, diagnostic imaging/procedures and pathology. See Table 2 for actual imaging/procedures and pathology costs incurred
§ The total cost of the Royal Flying Doctors Service inter-facility transfer flight from Horn Island to Cairns was \$8,730 which was funded by ARDMB through Queensland Department of Health. ARDMB recovered a proportion (72%) of the full amount from TCHHS.
‡ Rescue 700 flight from Boigu Island to Thursday Island and Thursday Island to Horn Island Airport
†† Wages of Queensland Ambulance Service paramedic to travel from Thursday Island to Boigu Island return on the Rescue 700 flight
¶¶ Includes road transfers from Cairns Airport to Cairns Hospital and road transfers and commercial flights from Cairns to Boigu Island to repatriate

### Costs rounded to the nearest Australian dollar

The median length of stay in the Australian healthcare system for 19 PNG patients diagnosed with TB and medically evacuated between 2016 and 2019 was 57 days (26-107). The length of stay for the patient in the case study was 24 days. Patients who received care in both Thursday Island and Cairns Hospitals experienced a median length of stay of 102 days, compared to a median stay of 43 days when receiving care in both Thursday Island and Townsville Hospitals.

## Discussion

The economic burden of managing and treating TB patients is clear across different populations, and comparison of previously reported costs, with the cost of managing PNG nationals at the Australian border is complex. The cost to medically evacuate and manage a patient with pulmonary TB in this study was AUD 124,280. This is more than ten-fold higher than the average cost of providing full treatment to fully susceptible TB patients in Victoria, Australia, at AUD 11,583 [17]. Similarly, meta-analyses of TB treatment costs in the European Union estimated the average total cost to treat a fully susceptible TB patient was EU 10,282 (AUD 17,213.47) [18]. It is important to point out that this patient had drug-susceptible TB, which is associated with greatly reduced in-patient care expense compared to patients with highly drug-resistant TB. Most published cost analyses report the total cost of administering a full course of anti-TB treatment, whereas the primary goal for managing a critically unwell PNG national TB patient entering Australia via the TSPZ is to stabilise, diagnose, commence on treatment and refer back to the PNG health system for continuation of their treatment and care.

In total, TB-related presentations make up a small proportion of all patients who receive and are retrieved for care in the Torres Strait. In a typical year, there are just under 100 aeromedical evacuations from Torres Strait outer islands involving PNG nationals, of which around ten are TB-related. The designated funds provided by the Australian Government Department of Health to compensate Queensland Health for healthcare provision to PNG nationals seeking healthcare via the TSPZ under Schedule B is AUD 4.7million per year. If each aeromedical evacuation costs a similar amount to our case study, there is a total shortfall in funding of over AUD 7.5 million per year to medically evacuate and manage PNG nationals. However, the true costs are unknown as analyses such as in this study are not routinely performed. In addition to better compensation for Queensland Health, greater transparency of expenditure and contractual milestone attainment could be achieved if the funds were held separately instead of being absorbed into HHSs.

As a result of the COVID-19 pandemic-related border closure between Australia and PNG in March 2020, residents of the Treaty villages living adjacent to the Australian border have experienced a reduction in access to TB diagnostics, treatment and critical care, and it is likely that household transmission of TB has increased during this time. As Australia prepares to open up to the world once more, it will be imperative that Queensland Health are prepared for a potential influx of patients with critical care needs from PNG.

## Conclusion

Funding of emergency evacuations for TB and other illnesses in this remote region, is necessary, unavoidable and expensive, and current levels of funding do not meet the demand. The high cost of patient care in such a remote setting, points to the benefit of supporting patients with critical care needs to prevent onward transmission of TB, as well as adequate budget allocation for aeromedical evacuation when required.

## Data Availability

Data are available from Queensland Health upon request, subject to Ethics approval and Public Health Act authorisation.

https://doi.org/10.6084/m9.figshare.16632823.v1

## Appendix A. Extended methodology: the cost to medically evacuate and manage one Papua New Guinea national diagnosed with pulmonary tuberculosis within the Australian health system.

The cost of this patient’s outpatient and inpatient clinic visits was obtained from Queensland Health’s fees and charges register [19]. Hospital and Health System cost categories included outpatient services confirmed as a Category 2 Emergency, pharmaceuticals, inpatient hospital stay, Royal Flying Doctors Service inter-facility transfer flight and other travel. The cost of inpatient hospital admission for Medicare ineligible patients includes clinician wages, meals, cleaning, pathology / procedures and diagnostic imaging.

Outpatient service expenses provided at Boigu Island Primary Health Centre include wages, cleaning, diagnostic imaging/procedures and pathology. Pharmaceuticals were calculated separately. Wages for the Torres and Cape TB Control Unit staff involved in providing an outpatient service to manage this patient were also calculated and reported separately.

Most travel-related costs (excluding aeromedical retrieval within the Torres Strait Islands) are borne by the Hospital and Health System in the jurisdiction where the patient first presented, regardless if additional travel across Queensland was required. All costs were reported separately for each health facility and the total cost of each category was provided.

The costs of pharmaceuticals were calculated by determining the full cost for any one item (i.e. box of Isoniazid 300mgs tablets), and multiplying this cost by the number of boxes or fractions of boxes required (this was obtained by dividing the number of doses administered to the patient by the total number of doses per box). Pathology was calculated per pathology request form rather than per test ordered, as per State billing requirements [20]. Item numbers for imaging services provided were obtained from the Queensland Health Medicare Benefits Schedule (MBS) Radiology Billing Manual [21] and then cross matched for cost from the MBS online [22].

Travel costs (excluding Boigu Island to Thursday Island; Thursday Island to Horn Island and Horn Island to Cairns) were sourced from the Torres and Cape Hospital and Health System patient travel and finance/revenue teams. These travel costs pertain to commercial flights and patient transport to and from relevant airports and hospitals. Travel costs did not include any expenses incurred by the patient or the Western Provincial Health Authority in PNG.

## Acknowledgments

This work was supported by the Torres and Cape Hospital and Health Service Revenue Manager located on Thursday Island. The authors are grateful to the Queensland Aeromedical Retrieval and Disaster Management Branch and the Queensland Ambulance Service for access to data and revision of relevant sections within the manuscript.

## Footnotes

1 Traditional inhabitants of the Torres Strait Protected Zone and Treaty villages may cross this international border without passport or visa. DOI. https://doi.org/10.6084/m9.figshare.16632823.v1

